# Ability of calculated right adrenal vein aldosterone levels to identify the aldosterone-overproducing side in patients with primary aldosteronism undergoing adrenal venous sampling

**DOI:** 10.1101/2023.11.13.23298491

**Authors:** Lalita Tuandam, Supamai Soonthornpun

**Author notes:** **Corresponding author:** Supamai Soonthornpun, M.D., Department of Medicine, Faculty of Medicine, Prince of Songkla University, Hat Yai, Songkhla 90110 THAILAND.

## Abstract

**Backgroud:** Adrenal venous sampling (AVS) is a gold standard procedure to determine the dominant side of aldosterone secretion in patients with primary aldosteronism. Unsuccessful cannulation of right adrenal vein (RAV) is a common problem in performing AVS.

**Objective:** To use calculated aldosterone concentration in the RAV (cAldo_RAV_) for identifying the dominant side of aldosterone secretion.

**Design:** Retrospective study, 2011-2023.

**Methods:** Based on the assumption that cortisol production from both adrenal glands is equal, aldosterone concentration in the RAV was calculated by using the data from left adrenal vein (LAV) and inferior vena cava. The aldosterone concentration in the LAV (Aldo_LAV_) compared to the cAldo_RAV_ (Aldo_LAV_:cAldo_RAV_ ratio) was then used to determine the dominant side of aldosterone secretion in patients with primary aldosteronism.

**Results:** Of 117 patients with successful AVS, 95 (81.2%) had concordant results between adrenal imaging and AVS study and were used as the gold standard for studying diagnostic performance. The Aldo_LAV_:cAldo_RAV_ ratio with the cutoff values of ≥3 and ≤0.33 could identify unilateral diseases (left-sided and right-sided disease, respectively) with 93.8% sensitivity and 100% specificity. In 22 patients who had discordant results between adrenal imaging and standard AVS interpretation, 11 had concordant results when using the Aldo_LAV_:cAldo_RAV_ ratio.

**Conclusions:** The Aldo_LAV_:cAldo_RAV_ ratio can determine the dominant side of aldosterone secretion with high sensitivity and specificity. It can not only be used for patients with unsuccessful cannulation of RAV but also increase the concordance rate in those who have discordance between adrenal imaging and standard AVS interpretation.

## Introduction

Primary aldosteronism is a leading cause of secondary hypertension. Aldosterone-producing adenoma (APA) and idiopathic hyperaldosteronism (IHA) are major subtypes that account for about 95% of patients with primary aldosteronism.^1^ Generally, APA is unilateral and IHA is bilateral disease, but this is not always true. Accurate identification of the dominant side of aldosterone secretion in patients with primary aldosteronism is an essential step because the bilateral disease needs life-long treatment with mineralocorticoid receptor antagonists, while the unilateral disease can be cured with a unilateral adrenalectomy. Despite a computed tomography being an accurate imaging tool for evaluating adrenal glands, it still has a limitation for discriminating unilateral from bilateral disease.^2-4^ Adrenal venous sampling (AVS) is accepted as a gold standard procedure to determine the dominant side of aldosterone secretion in patients with primary aldosteronism. A common problem in performing AVS is an unsuccessful cannulation of the right adrenal vein (RAV) due to its anatomy, namely small diameter, short length and high variants. Pasternak et al. used the aldosterone:cortisol ratio in the left adrenal vein (LAV) compared to that in the inferior vena cava (IVC) (LAV/IVC index) to determine the dominant side of aldosterone secretion.^5^ This index had a high specificity but modest sensitivity for determining unilateral disease. In addition, other studies have found that the cutoff value and diagnostic performance of this index were inconsistent.^6-12^

Based on the assumption that the cortisol production from both adrenal glands is equal, the aldosterone concentration in the RAV (Aldo_RAV_) can be calculated by using the data from the LAV and the IVC. The ratio between the aldosterone concentration in the LAV (Aldo_LAV_) and the calculated aldosterone concentration in the RAV (cAldo_RAV_) (Aldo_LAV_:cAldo_RAV_ ratio) was then used to determine the dominant side of aldosterone secretion.

## Materials and methods

This is a retrospective study of patients with confirmed or suspected primary aldosteronism who underwent AVS at Songklanagarind Hospital during the years 2011 to 2023. The study protocol was approved by the Ethics Committee of the Faculty of Medicine, Prince of Songkla University (REC.66-289-14-4). The diagnosis of primary aldosteronism was based on a saline infusion test. Confirmed diagnosis was made when plasma aldosterone concentrations at 4 h post-infusion were more than 10 ng/dL (277.4 pmol/L). The levels between 5-10 ng/dL (138.7-277.4 pmol/L) were considered as suspicious. Hypertensive patients with plasma aldosterone concentration >20 ng/dL (554.8 pmol/L), plasma renin activity <1 ug/L/h and spontaneous hypokalemia were also diagnosed as confirmed primary aldosteronism without performing saline infusion test.

Computed tomography of adrenal glands was done in all patients. The AVS with adrenocorticotropic hormone (ACTH) stimulation was performed in patients who were willing to undergo adrenalectomy if indicated. The ACTH was infused at the rate of 50 ug/h at least 30 min before and during the AVS. The AVS was performed by interventional vascular radiologists and was considered successful if cortisol ratio between adrenal vein and IVC (selectivity index) of both sides were more than 5. The dominant side of aldosterone secretion was determined by using the difference in aldosterone:cortisol ratio between RAV and LAV (lateralization index) of 3 times or more, while bilateral disease was diagnosed when the difference was less than 3 times. In case of failed cannulation of RAV only, the AVS was still considered successful if the difference in aldosterone:cortisol ratio between LAV and IVC (contralateral suppression index) was less than 0.5 and then the right adrenal gland was considered as the dominant side.^13^ Those with unsuccessful AVS were excluded from the study. Since it is difficult to confirm the dominant side of aldosterone secretion if the result of AVS is discordant with the adrenal imaging study, those who had concordant results between the AVS and imaging study were therefore selected to study the accuracy of using cAldo_RAV_ to identify the dominant side. Those with AVS results discordant with imaging study were subsequently analyzed for an agreement between using the Aldo_LAV_:cAldo_RAV_ ratio and standard AVS interpretation.

### Equation for cAldo_RAV_

The Aldo_RAV_ was calculated based on the assumption that the cortisol production from the right and left adrenal glands are equal. The aldosterone concentration in IVC (Aldo_IVC_) is equal to an average of the Aldo_LAV_ and the Aldo_RAV_, which is diluted by the blood flow in the IVC (dilution effect). The dilution effect was equal to cortisol concentration in the LAV (Cort_LAV_) divided by cortisol concentration in the IVC (Cort_IVC_), the so-called selectivity index of the left side. The Aldo_RAV_ was then calculated according to the equations shown below.

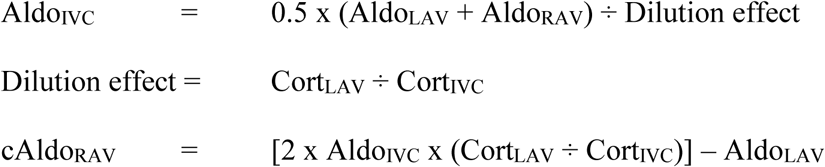

When the cAldo_RAV_ had a negative value, it was replaced by 100, which was the lowest value of aldosterone concentration in the non-dominant site found in our data. Finally, the dominant side of aldosterone secretion was determined by the Aldo_LAV_:cAldo_RAV_ ratio. Ratios of 3 or more indicated that left adrenal gland was the dominant side. Ratios of 0.33 or less indicated that right adrenal gland was the dominant side. Ratios between 0.33 and 3 indicated bilateral disease.

Plasma aldosterone concentration and plasma renin activity were measured by radioimmunoassay (DIASource ImmunoAssays, Louvain-la-Neuve, Belgium). Serum cortisol concentration was measured by electrochemiluminescent immunoassay (Roche Diagnostics, Mannheim, Germany)

### Statistical analysis

The diagnostic performance was determined by the area under the receiver operating characteristic (ROC) curve, sensitivity and specificity. The scatterplot was generated by MedCalc® Statistical Software version 20.215 (MedCalc Software Ltd, Ostend, Belgium).

## Results

Of 117 patients with successful AVS, 95 (81.2%) had concordant results between AVS and adrenal imaging study. Of those with concordant results, 81 were unilateral disease (38 were left-sided and 43 were right-sided) and 14 were bilateral disease. The dominant side was determined by using the lateralization index in 80 cases and using the contralateral suppression index in 15 cases. The areas under the ROC curve of the Aldo_LAV_:cAldo_RAV_ ratio for left-sided and right-sided diseases were 0.980 and 1.00, respectively. The scatterplot of the Aldo_LAV_:cAldo_RAV_ ratio in those with unilateral and bilateral disease is shown in Figure 1A. The cutoff values of ≥3 and ≤0.33 could identify the left-sided and the right-sided disease, respectively, with 93.8% sensitivity and 100% specificity. Likewise, the cutoff values between 3-0.33 could identify bilateral disease with 100% sensitivity and 93.8% specificity. Only 5 patients (5.3%) had the Aldo_LAV_:cAldo_RAV_ ratio discordant with the standard AVS interpretation. Figure 1B shows the scatterplot of the LAV/IVC index in the unilateral and bilateral disease. When using the LAV/IVC index with the cutoff values of ≥5.5 and ≤0.5 originally proposed by Pasternak et al., the sensitivity and specificity for diagnosis of the unilateral disease were 63.0% and 92.9%, respectively. In other words, the cutoff values between 0.5-5.5 for diagnosis of bilateral disease had 92.9% sensitivity and 63.0% specificity. Since the area under the ROC curve of the LAV/IVC index for left-sided and right-sided disease was also high (0.968 and 1.00, respectively), we identified the optimal cutoff values derived from our data as ≥1.5 and ≤0.6, which yielded a 93.8% sensitivity and 92.9% specificity for the diagnosis of unilateral disease.

**Figure 1.**
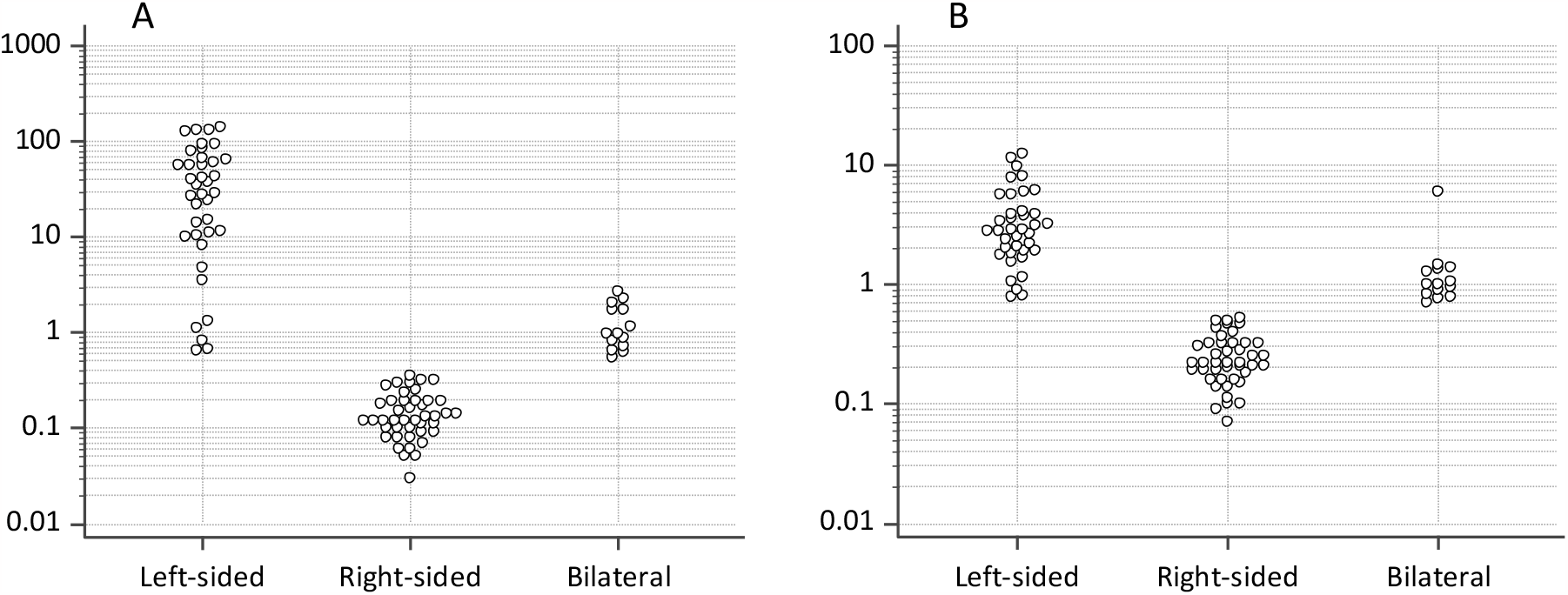
Logarithmic-scale scatterplot of the Aldo_LAV_:cAldo_RAV_ ratio (A) and the LAV/IVC index (B) according to the dominant side of aldosterone secretion.

Among the 22 patients who had discordant results between adrenal imaging and standard AVS interpretation, 20 were interpreted as the dominant side by using the lateralization index and 2 cases were interpreted using the contralateral suppression index. When using the Aldo_LAV_:cAldo_RAV_ ratio in these patients, the results were changed to concordance in 11 patients and the remaining half had the same results as the standard AVS interpretation (Table 1).

**Table 1.** The number of concordant and discordant pairs of adrenal imaging and AVS study using Aldo_LAV_:cAldo_RAV_ ratio in the 22 patients who had discordant results using standard AVS interpretation.

|  |  | Standard AVS interpretation |  |  | Aldo<sub>LAV</sub>:cAldo<sub>RAV</sub> ratio |  |  |
| --- | --- | --- | --- | --- | --- | --- | --- |
|  |  | Right-sided | Bilateral | Left-sided | Right-sided | Bilateral | Left-sided |
| Imaging study | Right-sided | 0 | 6 | 1 | 5 | 1 | 1 |
|  | Bilateral | 1 | 0 | 0 | 0 | 1 | 0 |
|  | Left-sided | 2 | 10 | 0 | 2 | 5 | 5 |
|  | Normal | 1 | 0 | 1 | 1 | 0 | 1 |

## Discussion

This is the first study that calculated the aldosterone concentration in the RAV by using a simple equation based on the assumption that each adrenal gland produces an equal amount of cortisol and used the Aldo_LAV_:cAldo_RAV_ ratio to identify the dominant side of aldosterone secretion in patients with confirmed or suspected primary aldosteronism. The Aldo_LAV_:cAldo_RAV_ ratio can diagnose unilateral and bilateral disease with high sensitivity and specificity comparable with the standard AVS interpretation using lateralization and contralateral suppression indices. This ratio also increased the concordance rate with imaging study in the patients who had discordant results when using the standard AVS interpretation.

The dilution effect in the equation was derived based on the assumption that each adrenal gland produces an equal amount of cortisol. If the cortisol production from the left adrenal gland was lower, the dilution effect was underestimated. On the contrary, if the cortisol production from the left adrenal gland was higher, the dilution effect was overestimated. Of 38 patients with left-sided disease, 5 had Aldo_LAV_:cAldo_RAV_ ratio less than 3. Theoretically, the falsely low Aldo_LAV_:cAldo_RAV_ ratio is most likely caused by overestimation of the dilution effect. However, as the cortisol concentrations in right and left adrenal veins in these patients were quite similar, the overestimation of the dilution effect could not explain the low Aldo_LAV_:cAldo_RAV_ ratio in these patients. An error in collecting blood samples and/or measuring aldosterone in the IVC may be another possible explanation for the falsely low Aldo_LAV_:cAldo_RAV_ ratio. The high sensitivity and specificity of the Aldo_LAV_:cAldo_RAV_ ratio for identifying the dominant side found in this study implies that the assumption of equal cortisol production from each adrenal gland is appropriate.

Apart from difficult cannulation and collection of blood samples, anatomical variants are also commonly found in the RAV.^14,15^ These could alter the AVS interpretation and mislead the results. On the other hand, LAV and IVC are easy to cannulate and draw blood. By using cAldo_RAV_ derived from the data obtained from LAV and IVC in this study, the discordance rate was reduced to 50% among those in whom standard AVS interpretation was discordant with imaging study, whereas the concordance rate was still very high (94.7%) in those who had concordant results between standard AVS interpretation and imaging study. This is one of the advantages of using the cAldo_RAV_ for dominant side determination in addition to those with unsuccessful cannulation of RAV.

Pasternak et al. originally used the LAV/IVC index to predict the dominant side of aldosterone secretion and found that the cutoff values of the LAV/IVC index of ≥5.5 and ≤0.5 predicted unilateral disease with 63% sensitivity and 100% specificity.^5^ These findings were similar to this study. However, when using the optimal cutoff values derived from our data of ≥1.5 and ≤0.6, it was found that the LAV/IVC index had a similar diagnostic performance to the Aldo_LAV_:cAldo_RAV_ ratio. An inconsistent cutoff value of the LAV/IVC index was also found in other studies.^8-11^ The cutoff values of the Aldo_LAV_:cAldo_RAV_ ratio of ≥3 and ≤0.33 means that left-sided disease is indicated when the Aldo_LAV_ is at least 3 times higher than the cAldo_RAV_. Conversely, right-sided disease is indicated when the Aldo_LAV_ is at least 3 times lower than the cAldo_RAV_. These cutoff values correspond with the criteria for a lateralization index of ≥3 used in this study. This is another advantage of using the Aldo_LAV_:cAldo_RAV_ ratio for dominant side determination.

There were some limitations in this study. First, the number of patients who had bilateral disease was small when compared to those with unilateral disease. The low number of bilateral disease in this study may be from a low sensitivity of saline infusion test to detect IHA.^16^ Second, the patients with discordant results between imaging and AVS study were treated with medications; the final diagnoses were therefore inconclusive. This is the reason why only those with concordant results were selected for analysis of diagnostic performance. Finally, it is the first time that the Aldo_LAV_:cAldo_RAV_ ratio has been proposed; validation studies are warranted to confirm the consistency of the cutoff values and results.

In conclusion, the Aldo_LAV_:cAldo_RAV_ ratio can determine the dominant side of aldosterone secretion with high sensitivity and specificity. It can not only be used for patients with unsuccessful cannulation of RAV but also increase the concordance rate in those who have discordance between adrenal imaging and standard AVS interpretation.

## Acknowledgments

We are very grateful to Alan F. Geater for proofreading the manuscript.

## Author Contributions

LT collected and analyzed the data and wrote the manuscript. SS designed the study, analyzed the data, and wrote the manuscript. All authors reviewed and edited the manuscript and approved the final version of the manuscript.

## Data Availability statement

The data are available from the corresponding author on reasonable request.

